# Environmental enteric dysfunction and small intestinal histomorphology of stunted children in Bangladesh

**DOI:** 10.1101/2022.05.09.22274879

**Authors:** Md. Shabab Hossain, S. M. Khodeza Nahar Begum, M Masudur Rahman, Mashud Parvez, Ramendra Nath Mazumder, Shafiqul Alam Sarker, Md. Mehedi Hasan, Shah Mohammad Fahim, Md. Amran Gazi, Subhasish Das, Mustafa Mahfuz, Tahmeed Ahmed

## Abstract

There is lack of information on the histological characteristics of the intestinal mucosa in Bangladeshi children, which is considered to be the traditional gold standard for diagnosis of environmental enteric dysfunction (EED). The purpose of the study was to evaluate the intestinal histological characteristics of stunted children aged between 12-18 months with possible EED. 110 children with chronic malnutrition (52 stunted with length-for-age Z score, LAZ<-2 and 58 at risk of stunting with LAZ <-1 to -2) from the Bangladesh Environmental Enteric Dysfunction (BEED) study protocol who underwent upper gastrointestinal (GI) endoscopy were selected for this study. To explore the association of EED with childhood stunting, upper GI endoscopy was done and the biopsy specimens were studied for histopathology. Villous height and crypt depth were measured and the presence and intensity of inflammatory infiltrates in the lamina propria was investigated. Bivariate analysis was performed to examine the relationship between stunting and histologic morphology. More than 90% children irrespective of nutritional status were diagnosed to have chronic non-specific duodenitis on histopathology. Half of the children from both groups had villous atrophy as well as crypt hyperplasia and lymphocytic infiltration was present in more than 90% children, irrespective of groups. However, no statistically significant difference was observed when compared between the groups. The prevalence of chronic non-specific duodenitis in Bangladeshi children, irrespective of nutritional status, was high. A significant number of these children had abnormal findings in intestinal histomorphology.

**Author Summary:** EED or environmental enteropathy, results in poor gut health and suboptimal child growth, and is considered to play a major role on childhood stunting in the tropics. Growth faltering due to decreased nutrient absorption as a result of alteration of small intestinal histological structure manifested as blunting or atrophy of intestinal villi, hyperplasia or elongation of crypts and infiltration of inflammatory cells in the lamina propria, has been demonstrated as the basic characteristics of EED. The traditional gold standard for the diagnosis of EED is intestinal biopsy and assessment of the histomorphological alterations. There is lack of information on the histological characteristics of the intestinal mucosa in Bangladeshi children. The purpose of the study was to evaluate the intestinal histological characteristics of stunted children aged between 12-18 months with possible EED. 110 children with chronic malnutrition who underwent upper gastrointestinal (GI) endoscopy were selected for this study and the biopsy specimens were studied for histopathology. Villous height and crypt depth were measured and the presence and intensity of inflammatory infiltrates in the lamina propria was investigated. More than 90% children irrespective of nutritional status were diagnosed to have chronic non-specific duodenitis. Half of the children from both groups had villous atrophy as well as crypt hyperplasia and lymphocytic infiltration was present in more than 90% children, irrespective of groups. However, no statistically significant difference was observed when compared between the groups.

## Introduction

Malnutrition accounts for almost half of all under-five child deaths ^1^, especially in lesser developed countries ^2^. Chronic malnutrition is frequently manifested as linear growth failure or stunting ^3^, affecting an estimated 150 million children below 5 years of age ^4^. About one-third of children in developing countries are stunted, accounting for 14–17% of mortality in under-five children ^5^. The survivors suffer from long-term sequelae including reduced neuro-developmental potential and poor cognitive function ^3^. According to a meta-analysis, each year two-hundred million children fail to achieve their growth potential as a result of stunting ^6^. Studies show that, children suffering from impaired linear growth during the first thousand days of life suffer from higher risk of chronic diseases and sub-optimal cognitive development causing sub-standard academic performance and financial productivity in the subsequent phases of life ^7^. Although Bangladesh has made substantial progress in terms of fighting malnutrition including stunting, with a reduction of stunting rates from 66% to 31% between the time period of 1990 and 2018, the prevalence rates of childhood stunting are still “very high” according to the criteria set by WHO ^8^.

There is high prevalence of chronic intestinal inflammation in the tropical parts of the world. The structural and functional abnormality of the child gut in this part of the world is also established ^5^. Certain changes in the small bowel function has been observed in the early phases of life in stunted children residing in developing countries, which is indicated by a change in the mucosal architecture of the intestine and chronic inflammation ^9^. These alterations appear to be caused because of prolonged exposure to a contaminated environment. However, the reversal of this condition has been observed in studies conducted earlier. These studies show that when a non-native person returns to their origins, even following a prolonged residence in a resource-poor setting, the gut alterations reverse back to normal. This led researchers to suggest the exposure to a contaminated environment to be responsible for such alterations ^9, 10, 11^, and the changes in mucosal architecture and intestinal inflammation is assumed to be the result of a prolonged exposure to enteric pathogens through fecal contamination, known as environmental enteric dysfunction (EED) ^9^.

EED, also recognized as environmental enteropathy or tropical enteropathy, results in poor gut health and suboptimal child growth ^12^, and is considered to play a major role on childhood stunting in the tropics ^3^. The mechanism of EED has been described earlier in many studies ^7, 13^. Growth faltering due to decreased nutrient absorption as a result of alteration of small intestinal histological structure manifested as blunting or atrophy of intestinal villi, hyperplasia or elongation of crypts and infiltration of inflammatory cells in the lamina propria, has been demonstrated as the basic characteristics of EED ^14^. EED is a clinically asymptomatic condition, and is only manifested visibly as growth impairment. The traditional gold standard for the diagnosis of EED is intestinal biopsy and assessment of the histomorphological alterations ^15^. It is evident that because of the invasive nature of the procedure of collecting intestinal biopsy samples, especially in young, apparently asymptomatic children, ^7^ the number of studies conducted to investigate this are very limited. The small number of studies that aimed to explore this have found blunting of the intestinal villi, increase in the depth of the crypts and infiltration of inflammatory cells in the lamina propria of intestinal biopsy samples as the key hallmarks of EED ^15^.

There is insufficient information on the intestinal histological morphology of stunted children with possible EED in Bangladesh. With an aim to mitigate this knowledge gap, we sought to explore these parameters in children aged between 12-18 months of age.

## Materials and methods

### Study site and Data collection

This study is a part of the Bangladesh Environmental Enteric Dysfunction (BEED) study which was conducted among the residents of *Bauniabadh* slum in Mirpur, Dhaka. BEED study was a community-based nutrition intervention study targeted at validating non-invasive biomarkers of EED with small intestinal biopsy samples and to have a better understanding of the disease pathogenesis. In this study, children aged between 12-18 months, who are stunted (length-for-age Z-scores (LAZ) <-2) and those who are at risk of stunting (LAZ <-1 to -2) were enrolled. The participants received on site nutritional intervention for 90 days consisting of one egg, 150 ml of whole milk and micronutrient sprinkles, as well as, nutritional counseling daily, six days a week. The details and overall design of the BEED study have already been published ^16^. Participants failing to respond to nutritional therapy and also negative for secondary malnutrition, i.e. tuberculosis, parasitic infection were considered as probable cases of EED and upper GI endoscopy was performed. For this particular study, a total of 110 children (52 stunted, 58 at risk of stunting) were enrolled from July 2016 to March 2019. Ethical approval was taken from Institutional review committee of icddr,b. The protocol number is PR-16007. A written informed consent for the intervention as well as the upper GI endoscopy and biopsy was obtained from the parents of children after explaining the aims and procedures of the study.

### Duodenal biopsy

Pediatric UGI endoscopy procedures were performed by expert endoscopists (MMR and MRB) at the Apollo Hospitals, Dhaka and Bangladesh Specialized Hospital, Dhaka using Olympus GIF Type Q180Z scope under general anesthesia. Biopsy specimens were collected from the duodenal bulb using Radial Jaw™ 4 Pediatric 2.0 mm single use biopsy forceps (Boston Scientific Corporation, Marlborough, USA) and were kept in 10% buffered formalin solution containing vials for the purpose of fixation. Paraffin sections were prepared and stained by hematoxylin and eosin. All biopsies were examined by expert pathologists (KNB and MP), who were absolutely blinded to the case histories. Estimation of the villous height, crypt depth and presence and intensity of inflammatory infiltrates in lamina propria as well as counting of the intraepithelial lymphocytes (IELs) was done ^17^.

### Definition of duodenitis and evaluation of intestinal histo-morphology

The definitions and classification of duodenitis used in this study have already been published elsewhere ^18^. Non-specific duodenitis was defined as the inflammation and morphological alterations of the duodenal mucosa, not known to be associated with any other pathology ^19^. Specific or secondary duodenitis was defined as the presence of inflammation and similar alterations of the duodenal mucosa due to a disease; for instance, Crohn’s disease, sarcoidosis, etc ^20^. Whether the duodenitis was active (chronic active duodenitis) or not was determined on the basis of the presence of neutrophilic infiltration or polymorphonuclear invasion, characterized by epithelial degeneration and regeneration with intercellular edema ^21^. Figure 1 shows the classification of duodenitis based on pathogenesis and cellular activity ^18^.

**Figure 1.**
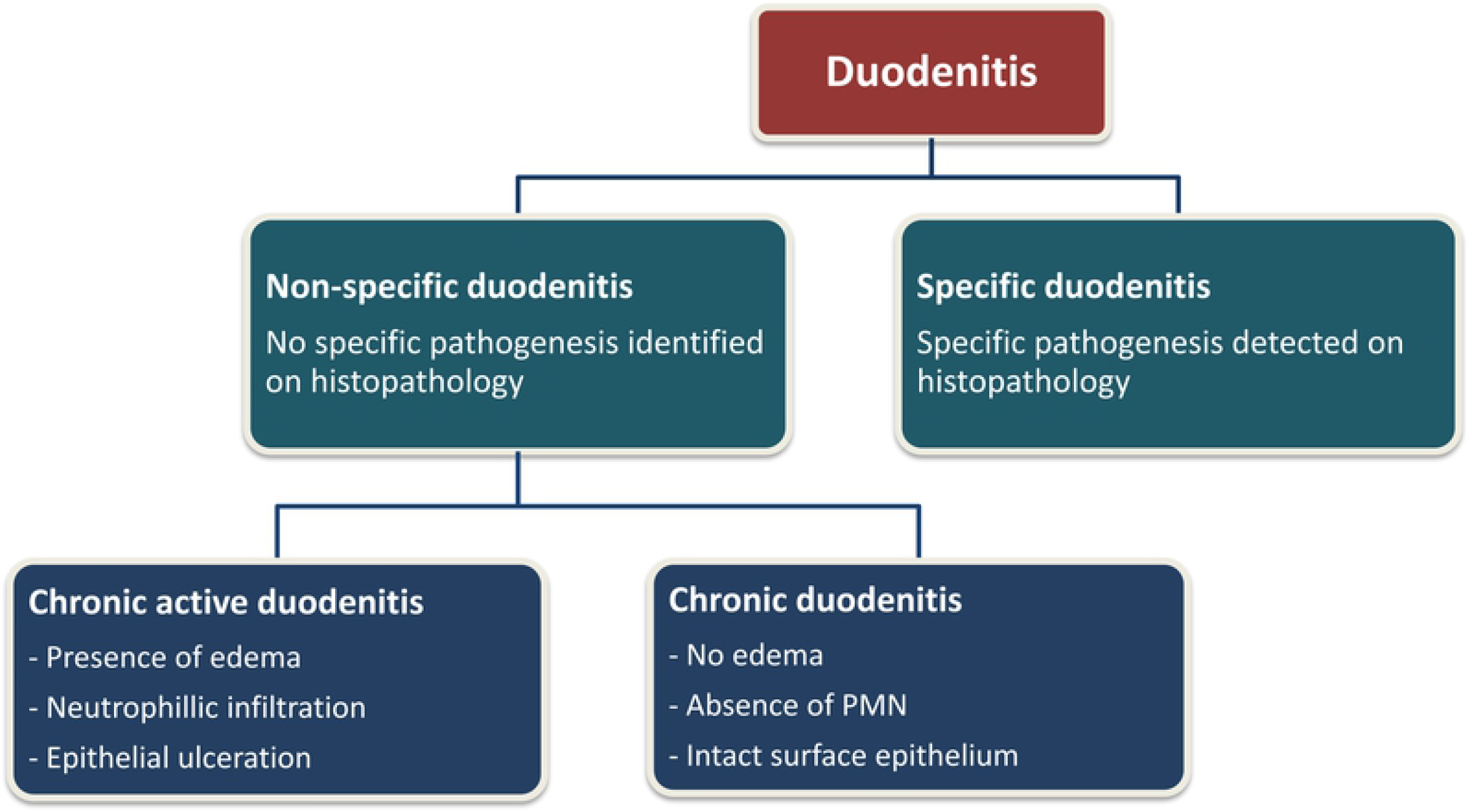
Classification of duodenitis based on pathogenesis and cellular activity

As reported in several studies ^5, 12, 13, 14, 22, 23^, morphometry of intestinal histology consisted of certain estimations related to remodeling of the crypts and villi with inflammatory infiltrations in the lamina propria. The schemes used to assess the morphological alterations in this study were also based on these parameters, which has already been published elsewhere ^18^. Shortening of the villous height was indicated by flattening of the mucosal surface secondary to the atrophy of the villi and was referred to as villous atrophy or blunting. Crypt changes consisted of elongation of the crypts or hyperplastic crypts ^24^. Complete flattening of the villi with a V:C ratio varying from 0:1 to 1:1 was defined as total villous atrophy, while partial blunting of the villi and a mild reduction of the villous height than normal, respectively, were referred to as subtotal and mild villous atrophy ^24^.

The lamina propria usually consists of plasma cells and lymphocytes, even in absence of active inflammation ^24^. Slides having an obvious presence of excess inflammatory cellular infiltrates than normal as estimated by expert eye were considered. The presence and intensity of inflammatory cellular infiltrates was further divided into marked cellular infiltration, moderate cellular infiltration and mild cellular infiltration through microscopic exploration ^18^. An intense and diffuse inflammatory infiltration that was clearly distinguishable on naked eye was referred to as marked cellular infiltration while moderate cellular infiltration was defined as the presence of a lymphoid aggregation or follicle. A higher number of infiltrates than normal was defined as mild cellular infiltration. Figure 2 shows the schematic representation used for the assessment of the morphological changes described above ^18^.

**Figure 2.**
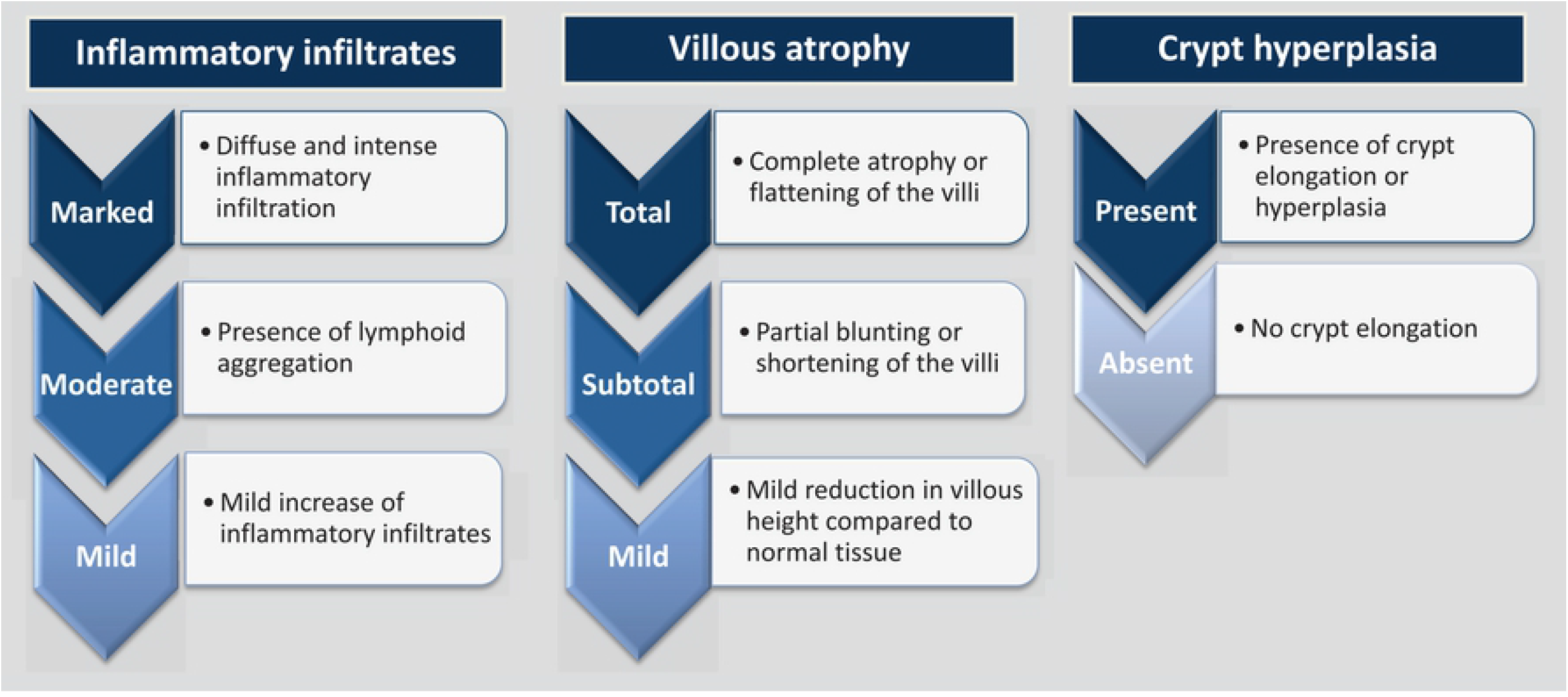
Schematic presentation for assessment of the histological alterations of intestinal mucosa

Increased numbers of IELs in an otherwise normal duodenal biopsy sample usually indicates certain immunological conditions; such as, lymphocytic/collagenous colitis, bacterial overgrowth and gluten sensitive enteropathy. Above 30 IELs/100 enterocytes was considered as intraepithelial lymphocytosis. IELs were estimated by counting the number of lymphocytes per 20 enterocytes present on a random villous tip, following the summation of the numbers of these lymphocytes from such 5 random villi ^24^. 20 enterocytes multiplied by 5 villi resulted in the number of lymphocytes per 100 enterocytes. Subjective morphologic analysis of the mucosal surface architecture was done using LEICA DM 1000 LED microscope.

### Statistical analyses

Mean values, standard deviation (SD) and 95% confidence intervals (CI) of means were used to describe the distribution and prevalence and bivariate analysis was performed to explore the relation between malnutrition and the histological features. Pearson’s chi-square test or Fisher’s exact test, whichever applicable, was used to compare the distribution of duodenitis and the histological features between the stunted and at risk of stunting children. Statistical significance was defined as a p-value of less than 0.05. Statistical analyses were performed using SPSS version 20.0 (IBM).

## Results

110 children (52 stunted, 58 at risk of stunting) with mean age of 18±2 months underwent upper GI endoscopy. Among them, 64 (58.2%) children were female. Representative histological images of normal villous architecture, mild villous atrophy, subtotal villous atrophy and total villous atrophy with crypt hyperplasia obtained using hematoxylin and eosin (H&E) stain are shown in Fig. 3 a, b, c and d respectively.

**Figure 3.**
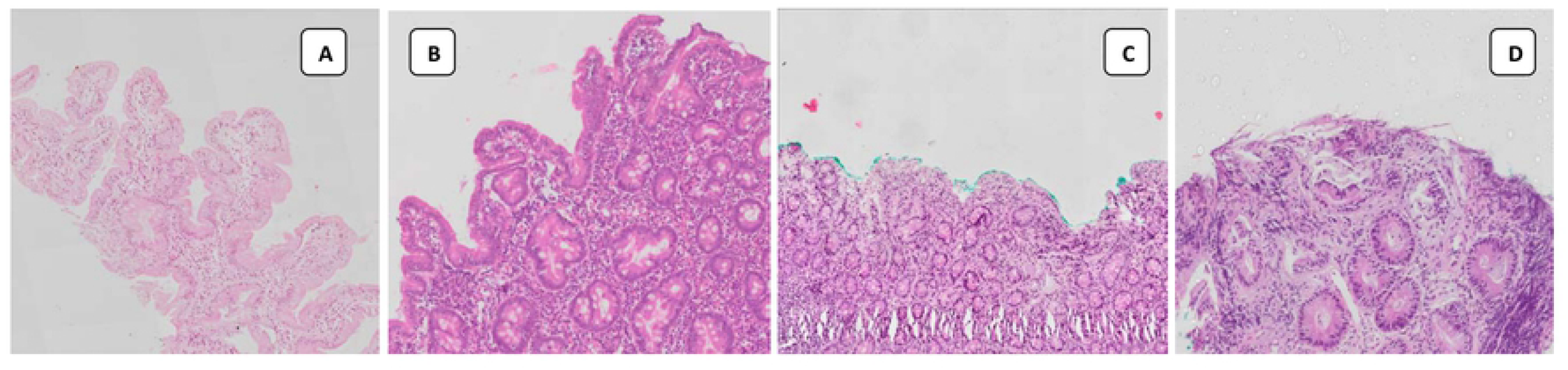
Representative histological images of villous height obtained using hematoxylin and eosin (H&E) stain

On histopathology, most of the children from both stunted and at risk of stunting groups suffered from chronic non-specific duodenitis. The proportion of chronic duodenitis and chronic active duodenitis in both groups were similar and no statistically significant difference existed between the groups. A small number of children belonging to both stunted and at risk of stunting cohorts were found to have abnormal intra-epithelial lymphocyte count in the biopsy samples. Presence of duodenitis did not differ significantly between the two groups of children. Table 1 shows the distribution of duodenitis in children.

**Table 1.** Types of duodenitis in children

| Duodenitis |  | Child Cohort |  |
| --- | --- | --- | --- |
|  | Stunted (†n= 52) | At risk (†n= 58) | †P-value |
| **Based on known pathology** |  |  |  |
| Normal | 3 (6%) | 3 (5%) | 1.00 |
| Specific | 0 (0%) | 0 (0%) | - |
| Non-specific | 49 (94%) | 55 (95%) | 0.62 |
| **Based on tissue morphology** |  |  |  |
| Normal | 3 (6%) | 3 (5%) | 1.00 |
| Chronic | 27(52%) | 26 (45%) | 0.46 |
| Chronic active | 22 (42%) | 29 (50%) | 0.42 |
| **Intraepithelial** | 2 (4%) | 3 (5%) | 1.00 |
| **lymphocytosis** |  |  |  |
| (†IEL>30) |  |  |  |

† n, Number of respondents; P-value, Significance level; IEL, Intraepithelial lymphocytosis

Morphology analysis shows, half of the children from both stunted and at risk of stunting groups had villous atrophy as well as crypt hyperplasia. Lymphocytic infiltration was present in more than 90% children, irrespective of groups. However, no statistically significant difference was observed between the groups. Figure 4 shows the morphologic distribution of histopathology findings.

**Figure 4.**
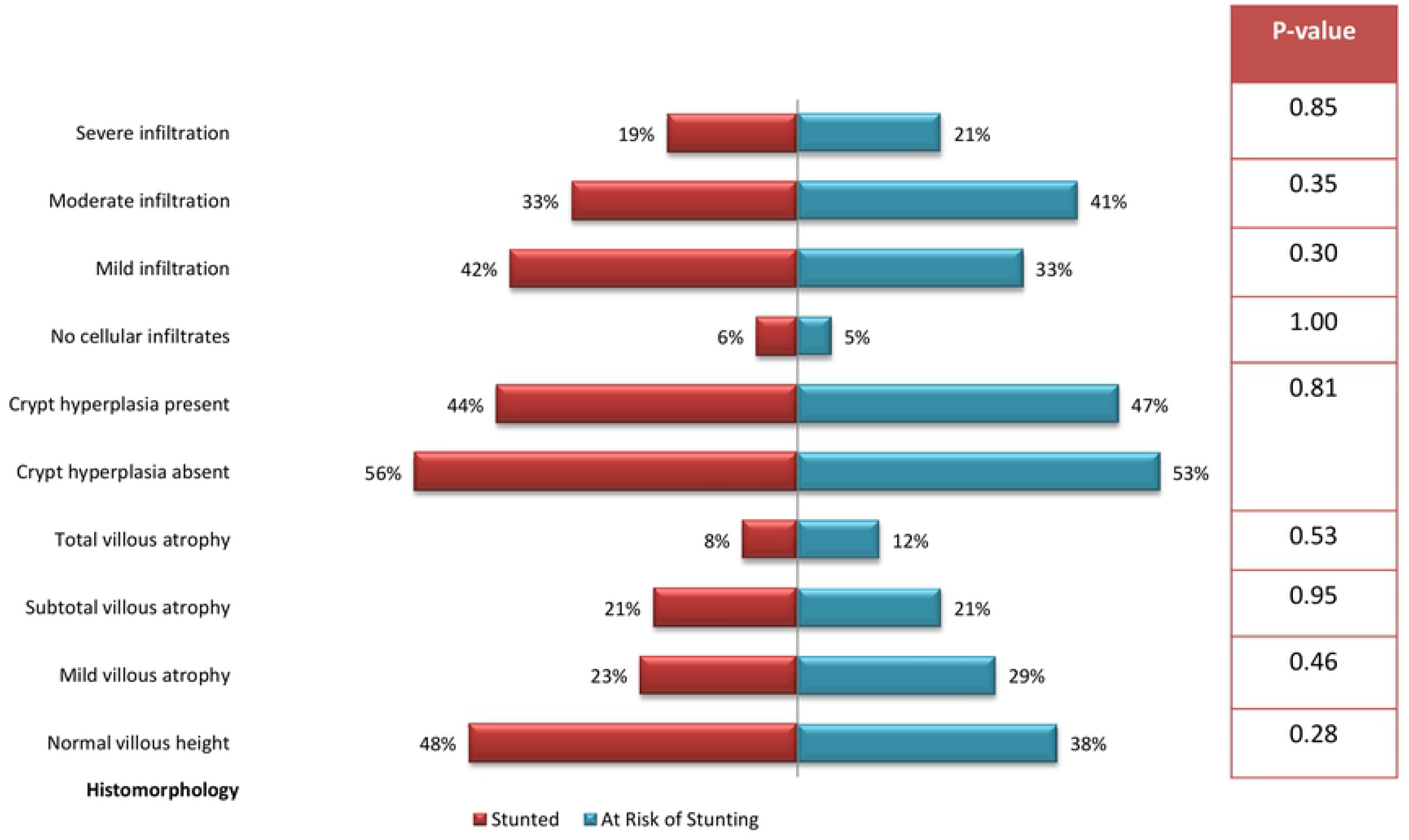
Histomorphological features of upper GI mucosa in children with stunting

## Discussion

This study showed a high prevalence of non-specific duodenitis on histopathology in children irrespective of their nutritional status. The finding is novel as after a thorough literature search, it was observed that there is paucity of data on the prevalence of duodenitis in this entire geographic region. Numerous studies were conducted to investigate pathologies of the stomach and esophagus ^25^, but only a few explored duodenal pathologies, especially in children ^25^. A similar study on malnourished adults has revealed very high prevalence of non-specific duodenitis based on histopathological evidence ^18^. A study aimed to explore *H. pylori* associated dyspepsia in children residing in urban slums has also shown a high prevalence of asymptomatic duodenitis ^26^. Studies based on esophagogastroduodenoscopy (EGD) procedures on US and Turkish children showed the prevalence of duodenitis to be 12.7% and 30.3%, respectively ^25, 27^. However, in contrast to stunted or at risk of stunting children in the present study, the children in those studies were relatively older, well-nourished and underwent EGD for diagnostic purpose. The commonest etiology responsible for duodenitis in studies conducted in Western countries was found to be celiac disease (CD), which accounted for 32% of the total cases ^28^. No children in the present study were found positive for celiac disease on serology ^29^ and the IEL count was also insignificant, which is an important histopathologic marker of CD ^30^. As the unique feature for all the children was only being stunted or at risk of stunting and they also did not have any pathology diagnosed by endoscopy, the high prevalence of duodenitis among these children was possibly related to EED and not due to celiac disease or any other GI disease ^16^. This also partly explains the high prevalence of the non-specific form of chronic duodenitis in the studied children, as the etiology could not be directly linked to any disease process. As the children underwent endoscopy to address the association of EED with stunting and not because of any known gastro-intestinal pathology, diagnosis of duodenitis with a specific etiology or specific duodenitis, would also be highly unlikely.

Approximately half of the children from both stunted and at risk of stunting cohorts had abnormal villous height as well as crypt depth. And lymphocytic infiltration was found to be present in majority of the participants, irrespective of groups. These findings are novel because this study is the first of its kind to explore the gut histomorphology in stunted children of this age group in Bangladesh. There are ethical challenges to obtain biopsy tissue from the intestine of young children without an evident clinical condition ^9^. As a result, a thorough literature search yielded very limited studies that have been conducted on asymptomatic children suffering solely from chronic malnutrition. A study done on 38 Gambian children suffering from protein energy malnutrition (PEM) had also shown that most of the children had villous atrophy or crypt elongation ^31^. Further studies aimed at obtaining serial biopsy samples at different ages and biopsy sites would be of great scientific value ^9^. No statistical significance was observed in terms of both duodenitis and intestinal histological morphology when both the groups were compared. As already mentioned, collection of small intestinal biopsy samples is an invasive procedure and hence is ethically and technically infeasible, particularly in young children without an evident clinical condition ^7^. As a result, biopsy samples from absolutely ‘healthy’ children could not be obtained. The comparison group here was at risk of stunting, and not a proper ‘healthy’ cohort, which could have yielded a better result.

From the morphological analysis it is evident that, nearly half of the children from both the groups had significant alterations in the intestinal histomorphology. As no other pathology which could have made a direct impact on these mucosal changes could be identified in these clinically asymptomatic children, EED may be a factor related with these findings. The children were provided with an intensive nutritional intervention prior to endoscopy as well as screened negative for other possible causes of malnutrition, yet, the children were stunted. Considering both the unexplained stunting and intestinal histomorphologic changes in the same children, EED might be the linking bridge and might also be a possible explanation. Thus, further studies associating EED markers with gut histomorphology in stunted children are warranted.

### Limitations

This study was conducted as a sub-study of the Bangladesh Environmental Enteric Dysfunction (BEED) study where the studied population of the parent study were only stunted (length-for-age Z-scores (LAZ) <-2) and at risk of stunting (LAZ <-1 to -2) children and therefore, by definition were not absolutely healthy. As collection of small intestinal biopsy samples in young children without an evident clinical condition is ethically infeasible, biopsy samples from absolutely ‘healthy’ children could not be obtained. Therefore, the comparison group was at risk of stunting, and not a proper ‘healthy’ cohort, which could have yielded a better result. Whether any association between EED markers with gut histomorphology exists or not was also not investigated, which also remained a limitation.

### Conclusion

Based on histopathology of the intestinal biopsy samples, chronic non-specific duodenitis was found to be very high in Bangladeshi children, irrespective of nutritional status. A significant number of these children had abnormal morphological findings in intestinal histopathology. As no other pathology could be identified in these children, EED may be a possible factor related to these findings, warranting further studies to establish this as a fact.

## Data Availability

The data underlying the results presented in the study are available from the corresponding author on reasonable request upon acceptance.

## Acknowledgements

This work was supported, in whole or in part, by the Bill & Melinda Gates Foundation [OPP1136751] and was funded by the Bill and Melinda Gates Foundation (BMGF) under its Global Health Program and the project investment ID is OPP1136751 (https://www.gatesfoundation.org/How-We-Work/Quick-Links/Grants-Database/Grants/2015/11/OPP1136751). Under the grant conditions of the Foundation, a Creative Commons Attribution 4.0 Generic License has already been assigned to the Author Accepted Manuscript version that might arise from this submission. icddr,b acknowledges with gratitude the commitment of BMGF to its research efforts. We express our sincere gratitude to our pediatric endoscopist, unfortunately who is no longer with us, late Prof. Dr. Md Muzibur Rahman Bhuiyan, former Senior Consultant, Department of Gastroenterology, Apollo Hospitals, Dhaka, Bangladesh. We express our sincere thanks to our colleagues at icddr,b and the study participants. icddr,b acknowledges with gratitude the commitment of the University of Virginia, Washington University in St. Louis, Dhaka Medical College and Hospital, Bangabandhu Sheikh Mujib Medical University, Dhaka Shishu Hospital, Bangladesh Specialised Hospital and Apollo Hospital, Dhaka to its research efforts. icddr,b is also grateful to the Governments of Bangladesh, Canada, Sweden and the UK for providing core/unrestricted support.

## Competing Interests

The authors declared no conflict of interest.

### Trial registration number

ClinicalTrials.gov ID: NCT02812615

(https://clinicaltrials.gov/ct2/results?cond=NCT02812615&term=&cntry=&state=&city=&dist=)

Date of first registration: 24/06/2016

